# Metabolic shift underlies recovery in reversible infantile respiratory chain deficiency

**DOI:** 10.1101/2020.04.21.20073759

**Authors:** Denisa Hathazi, Helen Griffin, Matthew J. Jennings, Michele Giunta, Christopher Powell, Sarah F. Pearce, Benjamin Munro, Wei Wei, Veronika Boczonadi, Joanna Poulton, Angela Pyle, Claudia Calabrese, Aurora Gomez-Duran, Ulrike Schara, Robert D.S. Pitceathly, Michael G. Hanna, Kairit Joost, Ana Cotta, Julia Filardi Paim, Monica Machado Navarro, Jennifer Duff, Andre Mattmann, Kristine Chapman, Serenella Servidei, Johanna Uusimaa, Andreas Roos, Vamsi Mootha, Michio Hirano, Mar Tulinius, Manta Giri, Eric P. Hoffmann, Hanns Lochmüller, Salvatore DiMauro, Michal Minczuk, Patrick F. Chinnery, Juliane S. Müller, Rita Horvath

## Abstract

Reversible infantile respiratory chain deficiency (RIRCD) is a rare mitochondrial myopathy leading to severe metabolic disturbances in infants, which recover spontaneously after 6 months of age. RIRCD is associated with the homoplasmic m.14674T>C mitochondrial DNA mutation, however only ∼1/100 carriers develop the disease. We studied 27 affected and 15 unaffected individuals from 19 families and found additional heterozygous mutations in nuclear genes interacting with mt-tRNA^Glu^ including *EARS2* and *TRMU* in the majority of affected individuals, but not in healthy carriers of m.14674T>C, supporting a digenic inheritance. The spontaneous recovery in infants with digenic mutations is modulated by changes in amino acid availability in a multi-step process. First, the integrated stress-response associated with increased *FGF21* and *GDF15* expression enhances catabolism via β-oxidation and the TCA cycle increasing the availability of amino acids. In the second phase mitochondrial biogenesis increases via mTOR activation, leading to improved mitochondrial translation and recovery. Similar mechanisms may explain the variable penetrance and tissue specificity of other mtDNA mutations and highlight the potential role of amino acids in improving mitochondrial disease.

## Introduction

Mitochondrial diseases are a large and clinically heterogeneous group of disorders that result from deficiencies in cellular energy production and affect at least 1 in 4300 of the population (Gorman,Chinnery et al., 2016). Although defective oxidative phosphorylation is the common pathway behind the disease, it is unknown why different mtDNA or nuclear mutations result in largely heterogeneous and often tissue specific clinical presentations. We have previously studied a rare and unique group of mitochondrial diseases, where life threatening symptoms present in infancy, but recover spontaneously after 6 months of age (infantile reversible mitochondrial diseases) (Boczonadi, Bansagi et al., 2015). The most common form of these conditions, reversible infantile respiratory chain deficiency (RIRCD OMIM *500009, previously called reversible infantile cytochrome c oxidase deficiency myopathy (Horvath, Kemp et al., 2009) is characterised by severe muscular hypotonia and weakness before 3 months of age (floppy baby), followed by a complete or almost complete spontaneous recovery after 6 months of age. Other organs are usually not involved. All patients affected by RIRCD carry the homoplasmic m.14674T>C/G mt-tRNA^Glu^ mutation, but only about a third of carriers within RIRCD families develop symptoms. Despite the striking reversible phenotype, less than 100 RIRCD patients have been described worldwide. The carrier frequency of the m.14674T>C variant is 0.01% in the healthy population (https://www.hmtdb.uniba.it/) and at a much higher frequency, 2/1,000 (0.2%) in Oceania, linked to the presence of this variant on the M27b1 mtDNA haplogroup, however no data are publicly available on the prevalence of RIRCD in Oceanian population (Duggan, Evans et al., 2014). The reasons for the markedly reduced penetrance are unknown.

A few other mitochondrial diseases have been also associated with a reversible infantile disease course (Boczonadi et al., 2015). Reversible infantile hepatopathy is caused by autosomal recessive mutations in the *TRMU* gene (tRNA 5-methylaminomethyl-2-thiouridylate methyltransferase, OMIM *610230) (Zeharia, Shaag et al., 2009), which uses cysteine to thiouridylate mt-tRNA^Glu^, mt-tRNA^Gln^ and mt-tRNA^Lys^. Cysteine is an essential amino acid in infants, due to the physiologically low activity of the cystathionine gamma-lyase (cystathionase) enzyme, and its low dietary availability in the first months of life contributes to reversible infantile hepatopathy (Sturman, Gaull et al., 1970, Zeharia et al., 2009). Also, heteroallelic *EARS2* (mitochondrial glutamic acid tRNA synthetase, OMIM*612799) mutations show partial recovery of neurological and muscle symptoms around 1 year of age in two third of patients (Steenweg, Ghezzi et al., 2012, Talim, Pyle et al., 2013).

In this study we utilized unbiased genomic sequencing, transcriptomic and proteomic approaches to explore the reasons for the markedly reduced penetrance and define the molecular mechanism of the reversibility in RIRCD associated with the homoplasmic m.14674T>C mutation.

## Results

### Clinical presentation

We studied 42 individuals (27 affected, 15 unaffected) from 19 families from 8 countries (UK, Germany, Sweden, Estonia, Italy, USA, Canada, Brazil) carrying the homoplasmic m.14674T>C mt-tRNA^Glu^ mutation (**Table 1, Supplementary Figure 1**). We present 10 new patients from 8 families, while 17 patients from 11 families were reported previously (Horvath et al., 2009, Houshmand, Larsson et al., 1994, Joost, Rodenburg et al., 2012, Uusimaa, Jungbluth et al., 2011). Informed consent was obtained from each participant approved by local ethics committees and the study was approved by the Yorkshire & The Humber - Leeds Bradford (13/YH/0310). The skeletal muscle and fibroblast samples were stored in the Newcastle Biobank of the MRC Centre for Neuromuscular Diseases (Reza, Cox et al., 2017).

**Table 1.**
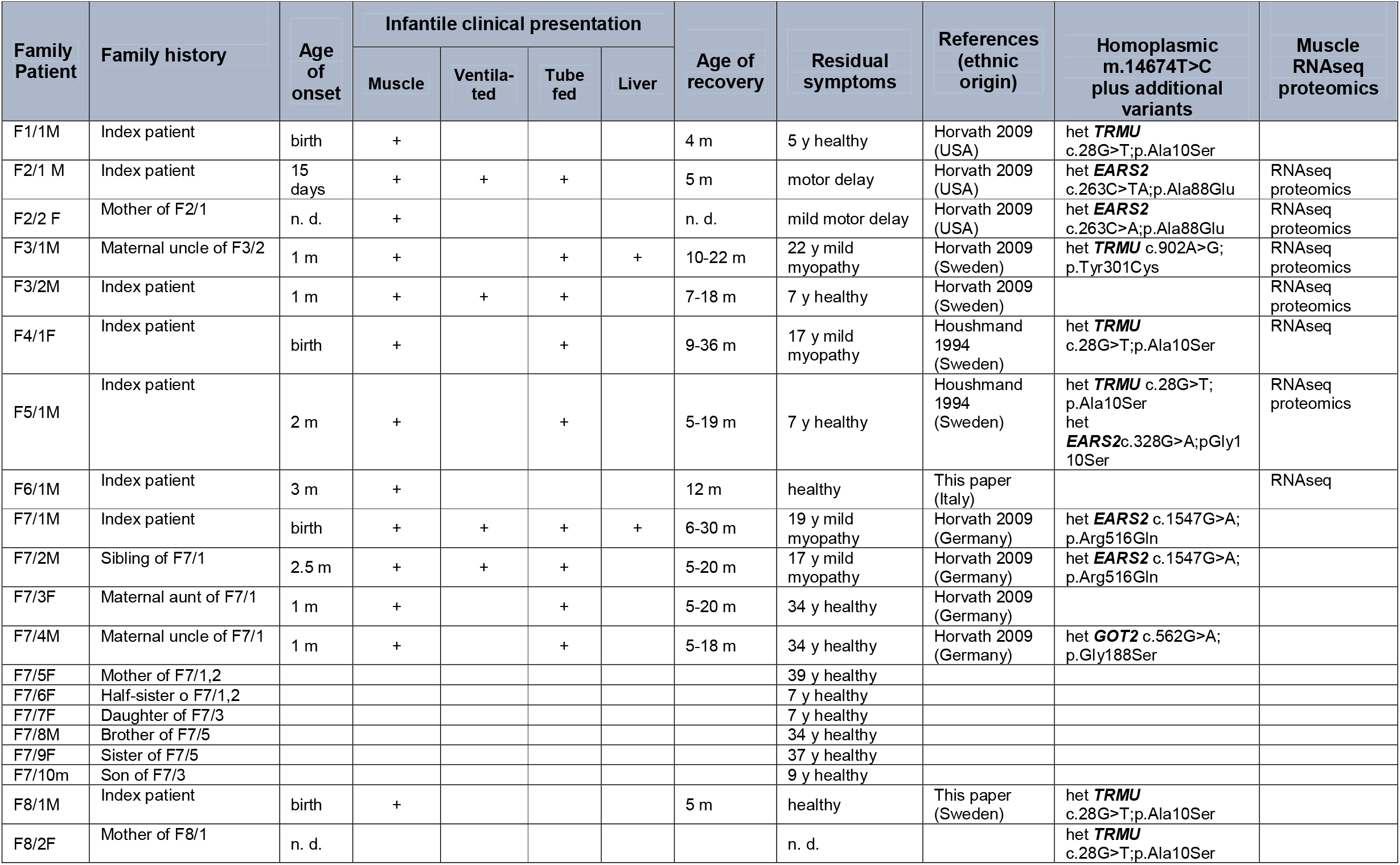
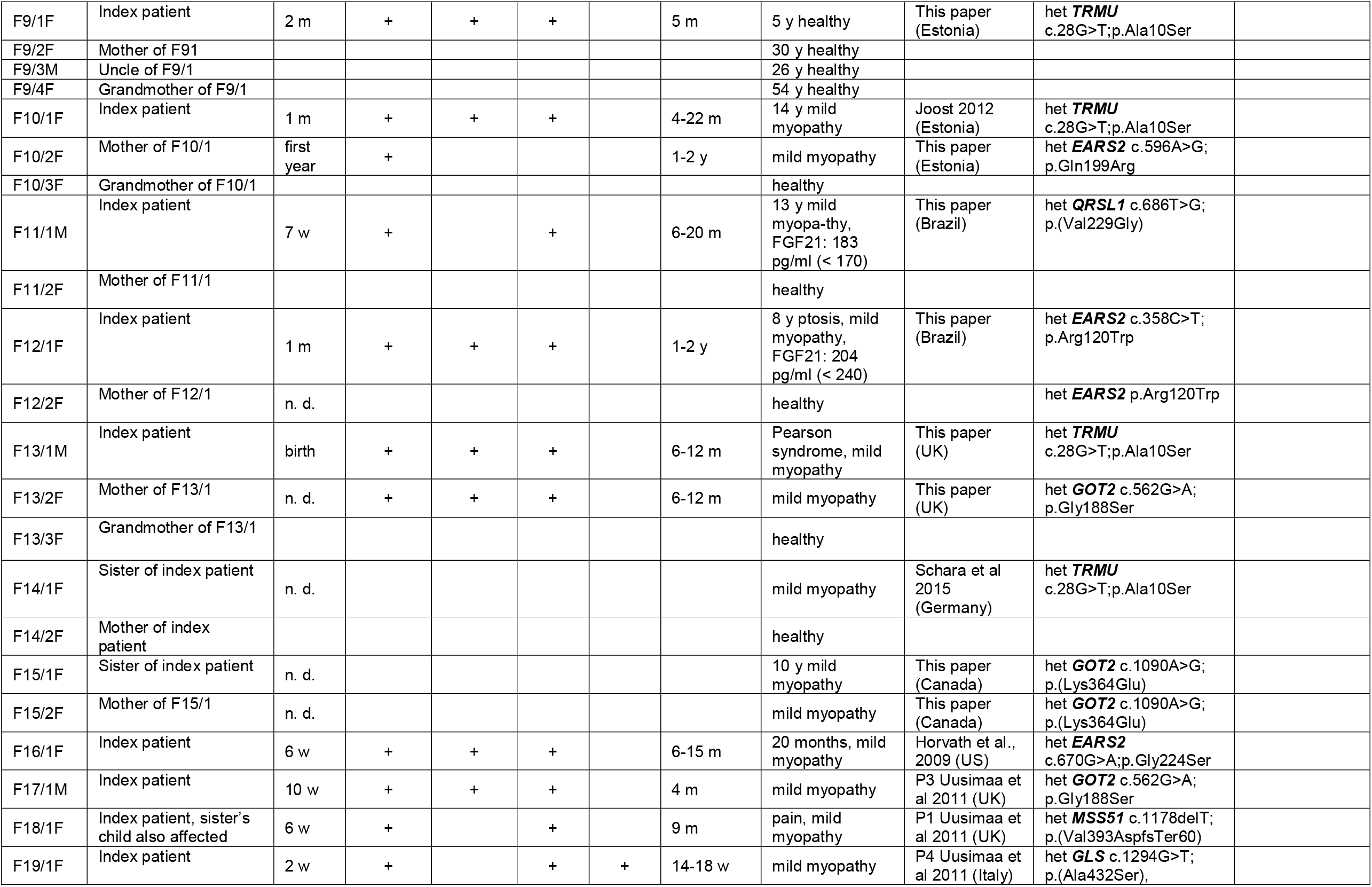
Summary of clinical presentations and genotypes of the index patients and their affected and unaffected family members in this study, all carrying the homoplasmic m.14674T>C mt-tRNA^Glu^ mutation

**Figure 1.**
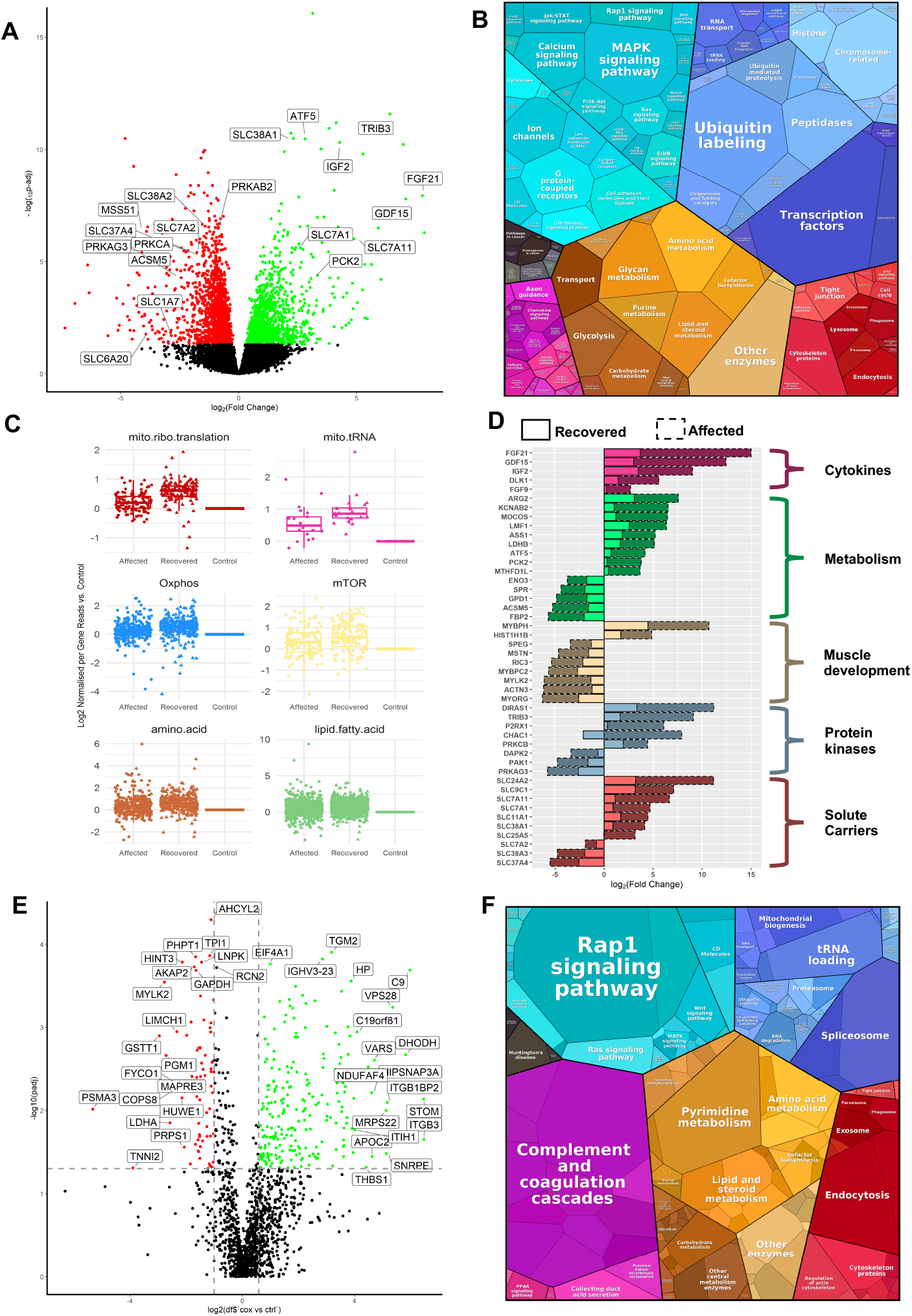
RNAseq differential expression (A-C) and Proteomics analysis (D & E) in human muscle. (A) Volcano plot shows genes where RNA reads are statistically significant under- or over-expressed in 6 RIRCD affected individuals versus 6 controls resulted from DESeq2 analysis. Decreased genes are represented in red while the up-regulated ones are in green while genes that were not statistically significant are represented in black. (B) Representation of the KEGG terms associated with differentially expressed RNAs and their corresponding proteins in affected patients versus control showing major pathway changes in patient muscle (https://bionic-vis.biologie.uni-greifswald.de/). (C) Abnormal transcription of genes involved in various biological metabolic pathways derived from our RNAseq data showing that their expression returned close to normal levels in patient muscle after recovery. (D) Relevant functional changes in gene expression showing the major proteins that return to baseline or contribute to the recovery in RIRCD patients. The data depicted is between the affected and recovered biopsies of the same patient (F6/1M). (E) Comparative proteome profiling of proteins that are significantly under- or over-expressed in RIRCD affected versus control individuals (p-Anova <0.05-statistically significant - horizontal line). Proteins which are decreased are represented in red while the up-regulated ones are in green while proteins that did not reach statistical significance threshold are marked in black. (F) Proteomap representation of the major altered pathways in RIRCD affected muscle compared to control individuals where the size of each circle or hexagon represents the fold change.

The clinical presentation of the patients and their family members homoplasmic for m.14674T>C is summarized in **Table 1**. In brief, 22 patients (13 male/9 female) presented with muscle weakness before 3 months of age and showed partial or complete recovery between 4-30 months of age (RIRCD), or a mild, residual myopathy in 10 individuals. An additional 4 female homoplasmic mutation carriers from 4 different families did not present with weakness in the first months of life, however developed mild non-progressive muscle weakness at a later age. Fifteen homoplasmic mutation carriers were healthy and never showed myopathy or any other symptom which can be associated with mitochondrial disease. Two generations were affected in 5 families, with siblings in 2 families.

### Whole exome sequencing (WES) to search for common genetic modifiers of m.14674T>C

WES was performed in 34 individuals (22 patients and 12 unaffected carriers). Mitochondrial DNA haplogroups were obtained from WES (Griffin, Pyle et al., 2014) and the homoplasmic m.14674T>C mutation was detected in all individuals on twelve different haplogroup backgrounds, demonstrating that this mutation has arisen independently in unrelated families **(Supplementary Figure 2, Supplementary Table 1)**. We did not identify any additional mtDNA variants in the affected individuals which could potentially affect mitochondrial translation and thus contribute to the clinical presentation.

**Figure 2.**
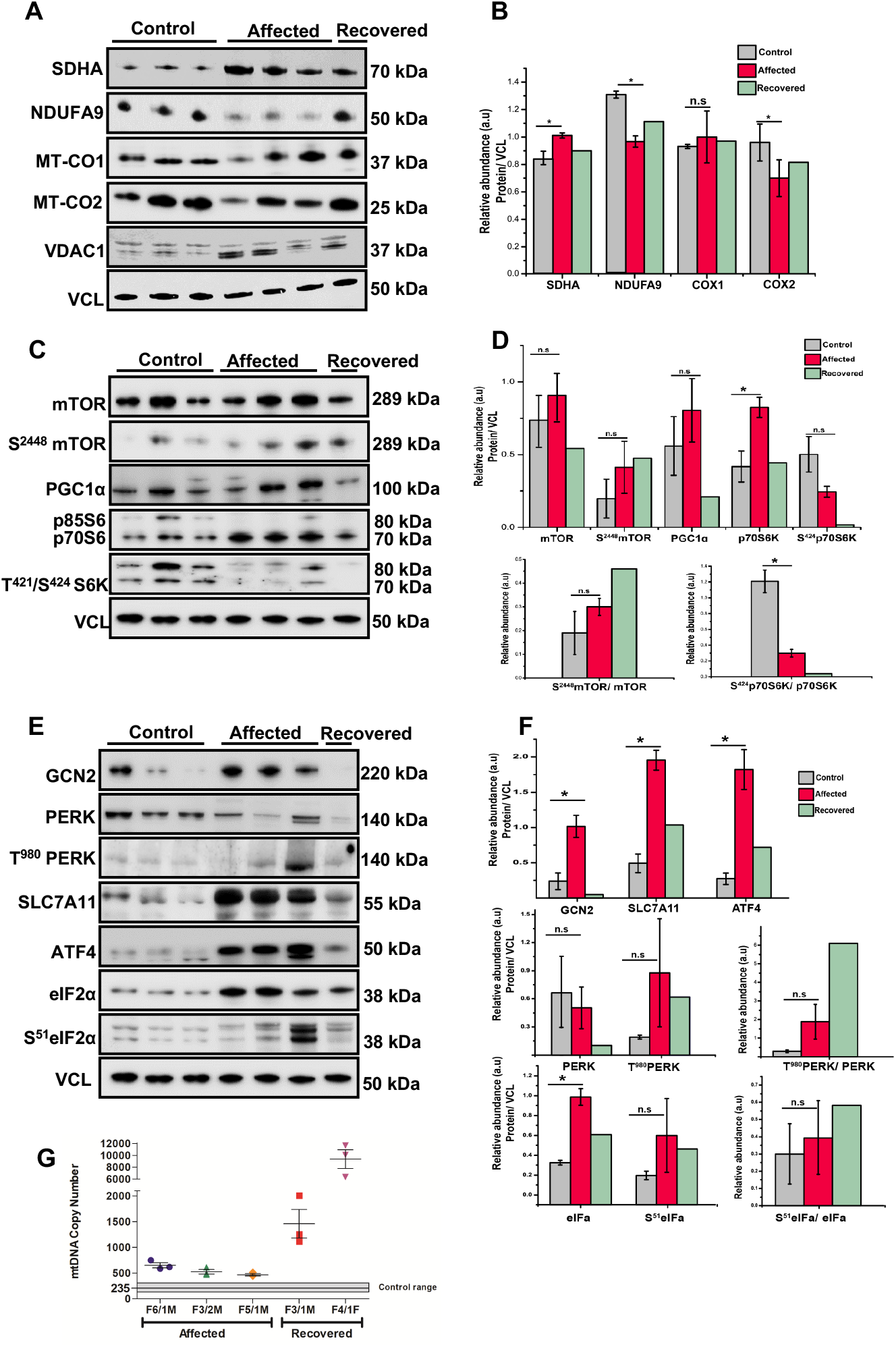
Immunoblotting in skeletal muscle of patients detected alterations of several mitochondrial complex subunits, the activation of integrated stress response and mTOR. (A) Western blotting analysis depicts mitochondrial proteins (NDUFA-Complex I, SDHA-Complex II, MTCO1-Complex IV and MTCO2-Complex IV) levels in control derived muscle (n=3), patient derived muscle during the symptomatic phase (n=3) and patient derived muscle after recovery (n=1). VCL was used as a loading control while the densitometry analysis was done based on the mitochondrial protein VDAC1. (B) Densitometry analysis of the immunoblotting from (A) showing that an increase in SDHA (Complex II) and MT-CO1(Complex IV) while NDUFA9 (Complex I) probably as a compensatory mechanism and MT-CO2 (Complex IV) show significant reductions as the majority of mitochondrial encoded proteins belong to this two complexes, thus being destabilized by the defective mitochondrial mistranslation. Graphs show means ± s.d of triplicate samples (controls and affected muscle) and results obtained from (n=1) recovered muscle. (C) Immunoblot analysis of mTOR pathway components: S^2448^mTOR, mTOR, PGC1α, T^421^/S^424^ P70S6K and P70S6K in control derived muscle (n=3), patient derived muscle during the symptomatic phase (n=3) and patient derived muscle after recovery (n=1). VCL was used as a loading control. (D) Densitometry analysis of the immunoblotting from (C) depicting an increase in mTOR in patient muscle consistent with an activation of downstream proteins such as PGC1α (increases mitochondrial biogenesis) and p70S6K (increases cellular proliferation). Graphs show means ± s.d of triplicate samples (controls and affected muscle) and results obtained from n=1 recovered muscle. Additionally to normalizing to VCL, phosphorylated proteins were also normalized to the total amount of non-phosphorylated counterpart in order to determine the correct phosphorylation amount with an increase in mTOR phosphorylation in recovered muscle compared to affected and control derived muscle. (E) Western blotting analysis of proteins involved in the amino-acid sensing and integrated stress response: GCN2, T^980^PERK, PERK, SLC7A11, ATF4, S^51^eIF2α and eIF2α in control derived muscle (n=3), patient derived muscle during the symptomatic phase (n=3) and patient derived muscle after recovery (n=1). All samples were normalized to VCL which was used as a loading control. (F) Densitometry analysis of proteins from (E) showing a significant increase of major transducers of ISR such as GCN2 and ATF4. Consistent with ISR activation is also the increase in SLC7A1. Graphs show means ± s.d of triplicate samples (controls and affected muscle) and results obtained from n=1 recovered muscle. In addition to normalizing to VCL, phosphorylated proteins were normalized to the total amount of non-phosphorylated protein. Similar results were observed in at least two independent results (due to the limited amount of sample). (*p<0.05; two-tailed one sample t-test). (G) Mitochondrial DNA copy numbers were determined in affected and recovered individuals and compared to control individuals using qPCR showing a significant increase in recovered muscle.

RIRCD manifested with ∼30% penetrance in the first year of life in previously reported families homoplasmic for m.14674T>C, raising the possibility of a genetic modifier (Boczonadi et al., 2015). We first hypothesized that a common genetic variant, either in affected patients or in healthy controls, may modify the clinical presentation of m.14674T>C(Boczonadi et al., 2015, Horvath et al., 2009) (**Supplementary Table 2**), however no single variant segregated in all affected or unaffected individuals. We identified a frequent (minor allele frequency or MAF 0.4723, homozygote frequency 0.11) missense variant c.67C>T, p.Arg23Trp in *PDE12* which was homozygous in 14 affected individuals from 8 out of 19 families (with different mtDNA haplotype), although three unaffected homozygous carriers were subsequently found in one family (Affection vs. *PDE12*, Fishers Exact p=0.087). PDE12 or 2’, 5’-phosphodiesterase 12 is a major factor for the quality control of mitochondrial non-coding RNAs and the lack of PDE12 results in a spurious polyadenylation of the 3’ ends of the mitochondrial (mt-) rRNA and mt-tRNA (Pearce, Rorbach et al., 2017). The c.67C>T, p.Arg23Trp variant however, did not affect the respiratory chain complexes and the processing of 3’ poly(A) tails of the 16S mt-rRNA, because introducing the mutant gene rescued the phenotype of *PDE12* knock-out cells (**Supplementary Figure 3)**.

**Figure 3.**
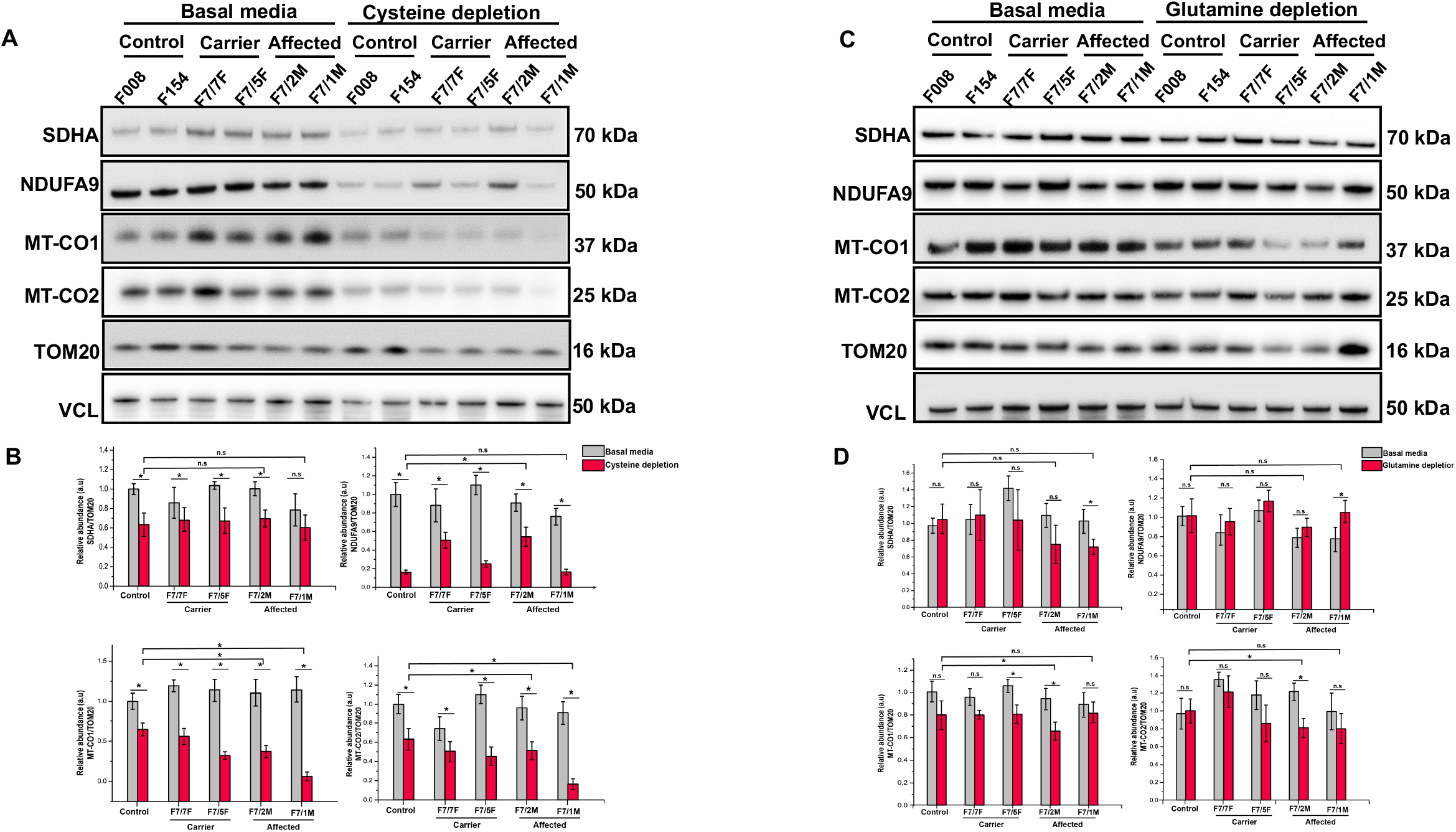
Cysteine depletion alters expression of complex I and IV in RIRCD fibroblasts. (A) Patient (F7/7F-healthy carrier, F7/2M-RIRCD, F7/5F-healthy carrier, F7/3F-RIRCD without a clear second mutation) and control fibroblasts were grown for 12 days in media with no added glutamine or glutamic acid and 5% dialyzed FBS. Proteins belonging to mitochondrial complexes (NDUFA-Complex I, SDHA-Complex II, MTCO1-Complex IV and MTCO2-Complex IV) were analysed via Western Blotting. VCL was used as a loading control and the densitometry analysis was done based on the mitochondrial protein VDAC1. Densitometry analysis of proteins from (A) showing no significant changes in OXPHOS proteins upon glutamine/ glutamic acid depletion. Graphs show means ± s.d of duplicate samples. (C) Patient (F7/7F-healthy carrier, F7/2M-RIRCD, F7/5F-healthy carrier, F7/3F-RIRCD without a clear second mutation) and control fibroblasts were grown for 12 days in media containing 0.02 mM cysteine and 5% dialyzed FBS. Cells were lysed and the above mentioned mitochondrial proteins were checked via Immunoblotting. VCL was used as a loading control while the densitometry analysis (D) was done based on the mitochondrial protein VDAC1. Although we could observe significant changes upon cysteine depletion in all conditions, the phenotype was exacerbated in the digenic fibroblasts compared to the controls. Graphs show means ± s.d of duplicate samples. All experiments were performed in triplicate n=3 and similar results were observed in all experiments (*P<0.05; two-tailed one sample t-test).

In 9 RIRCD patients from 8 families (with different mtDNA haplotype) we detected the heterozygous common c.28G>T; p.Ala10Ser *TRMU* variant **(Supplementary Figure 1, Supplementary Table 1**), which has been shown to result in reduced thiouridylation of mt-tRNA^Glu^ (Meng, Cang et al., 2017). It has a population total allele frequency of 0.09695 (gnomAD). Thus, the probability of carrying both p.Ala10Ser and m.14674T>C is approximately 1 in 100,000 (1×10^−5^), a frequency that is compatible with the rare occurrence of RIRCD in a subset of homoplasmic m.14674T>C carriers.

### Digenic inheritance of m.14674T>C with rare, damaging nuclear variants

Based on the frequency of m.14674T>C (0.01%) and the predicted 30% penetrance rate in previously reported families (Boczonadi et al., 2015), the estimated prevalence of RIRCD patients should be ∼1/30,000 (0.003%,) predicting >2,000 RIRCD patients only in the UK. However, less than 100 RIRCD patients have been reported world-wide to date, suggesting that at least 100 times more unaffected individuals and families carry the homoplasmic m.14674T>C change. This is supported by the existence of two m.14674T>C carriers in a Danish type 2 diabetes study (Li, Besenbacher et al., 2014) and two homoplasmic carriers in the NIHR BioResource Rare Diseases dataset. Heteroplasmic carriers (between 3-15% mutant allele) have also been found in three ‘1000Genomes’ samples and there are 11 carriers (9 females, 2 males) of the homoplasmic m.14674T>C in the UK Biobank European unrelated individuals (N = 358,916), confirming the MAF = 3×10^−5^ (Wei, Tuna et al., 2019). None of these carriers had any evidence of mitochondrial disease. The most likely scenario is that affected individuals carry another variant (or variants) in addition to m.14674T>C, and the additive effect of these variants may underlie the phenotype. Therefore, we filtered our WES data for damaging, rare variants specific to affected or unaffected individuals within the RIRCD pedigrees and detected 2,698 potentially damaging variants in affected and 1,067 variants in unaffected individuals within 19 families (**Supplementary Table 2)**. No homozygous or compound heterozygous pathogenic variants were detected in genes involved in mitochondrial translation in any of the affected individuals. However, 6 different rare heterozygous damaging variants in *EARS2*, the mt-tRNA synthetase responsible for aminoacylation of mt-tRNA^Glu^ co-segregated with the disease in 9 RIRCD patients from 6 families **(Supplementary Figure 1, Supplementary Table 1**).

Three *EARS2* variants identified in our families (c.328G>A; p.Gly110Ser, c.670G>A; p.Gly224Ser and c.1547G>A; p.Arg516Gln) have been previously reported as pathogenic mutations in compound heterozygous state with other mutations in patients with autosomal recessive leukoencephalopathy with brainstem and thalamus involvement and high lactate (LBTL, OMIM *612799) (Steenweg et al., 2012). The other 3 heterozygous *EARS2* variants (c.358C>T; p.Arg120Trp, c.596A>G; p.Gln199Arg and c.263C>T; p.Ala88Glu) were predicted to be damaging by multiple Annovar annotation databases and alter evolutionarily conserved amino acids, in known functional protein domains. The *EARS2* variants were all rare (MAFs<0.002) and no homozygous individuals are reported in control databases (ExAC, gnomAD) except for c.670G>A; p.Gly224Ser **(Supplementary Table 3**). Despite the recent publications on patients with recessive *EARS2* mutations, clinical symptoms have not been observed to date in heterozygous carriers of this gene (Steenweg et al., 2012). Based on the frequency of *EARS2* variants in international databases, we calculated that the cumulative frequency of carrying a heterozygous damaging variant in this gene is approximately 0.0135. Based on the 0.0001 MAF of m.14674T>C and 0.0135 cumulative MAF of heterozygous damaging variants in *EARS2* the probability to carry both is approximately 1 in a million (1.35×10^−6^). This number is compatible with the extremely rare occurrence of RIRCD and supports the observation in our 6 families that both mt-tRNA^Glu^ and *EARS2* variants contribute to the clinical manifestation of the disease.

A rare, damaging heterozygous *TRMU* variant (c.902A>G; p.Tyr301Cys) co-segregated with RIRCD in one family. It is also likely to contribute to the phenotype with a similar digenic mechanism. Additionally, a severely affected RIRCD patient carried heterozygous variants in both *EARS2* (c.328G>A; pGly110Ser) and *TRMU* (p.Ala10Ser) in addition to m.14674T>C, indicating a possible correlation between mutational load and disease severity.

Seven affected individuals without heterozygous variants in *EARS2* or *TRMU* carried heterozygous variants in human disease genes interacting with mt-tRNA^Glu^ such as *QRSL1* (another mt-tRNA synthetase aminoacylating mt-tRNA^Glu^), *GOT2* (mitochondrial glutamic-oxaloacetic transaminase) and *GLS* (mitochondrial glutaminase), both involved in glutamic acid metabolism (Friederich, Timal et al., 2018, van Karnebeek, Ramos et al., 2019, van Kuilenburg, Tarailo-Graovac et al., 2019). One patient carried a rare heterozygous frameshift variant in the gene *MSS51*, which has been shown to act as a translation activator of *MTCO1* and an assembly factor of cytochrome *c* oxidase (Garcia-Villegas, Camacho-Villasana et al., 2017, Moyer & Wagner, 2015), and had a significantly decreased gene expression in the affected patients’ muscle (family 18) (**Figure 1, Supplementary Table 3)**. Together the cumulative MAFs of damaging *EARS2, TRMU, QRSL1, GOT2, GLS* and *MSS51* variants found in the ExAC database (MAF=0.32) could explain the affected status of 32% of m.14674T>C mutation carriers in our cohort, which is consistent with previous observation that ∼30% of homoplasmic mutation carriers within RIRCD families develop the symptoms (Boczonadi et al., 2015). The cumulative incidence of damaging variants in *EARS2, TRMU, QRSL1, GOT2, GLS, MSS51* and m.14674T>C (**Supplementary Figure 4**) shows a significant increase in the mean number of variants in these genes between RIRCD affected and unaffected carriers (mean alleles: affected=2.29, unaffected=1.33; t=4.77; p=5.1×10^−5^) and even more between RIRCD and controls (mean alleles: affected=2.29, 1000 Genomes=0.73; t=10.89; p=3.1×10^−10^). None of the unaffected UK Biobank m.14674T>C carriers are heterozygous for pathogenic variants in *EARS2* or *TRMU*.

**Figure 4.**
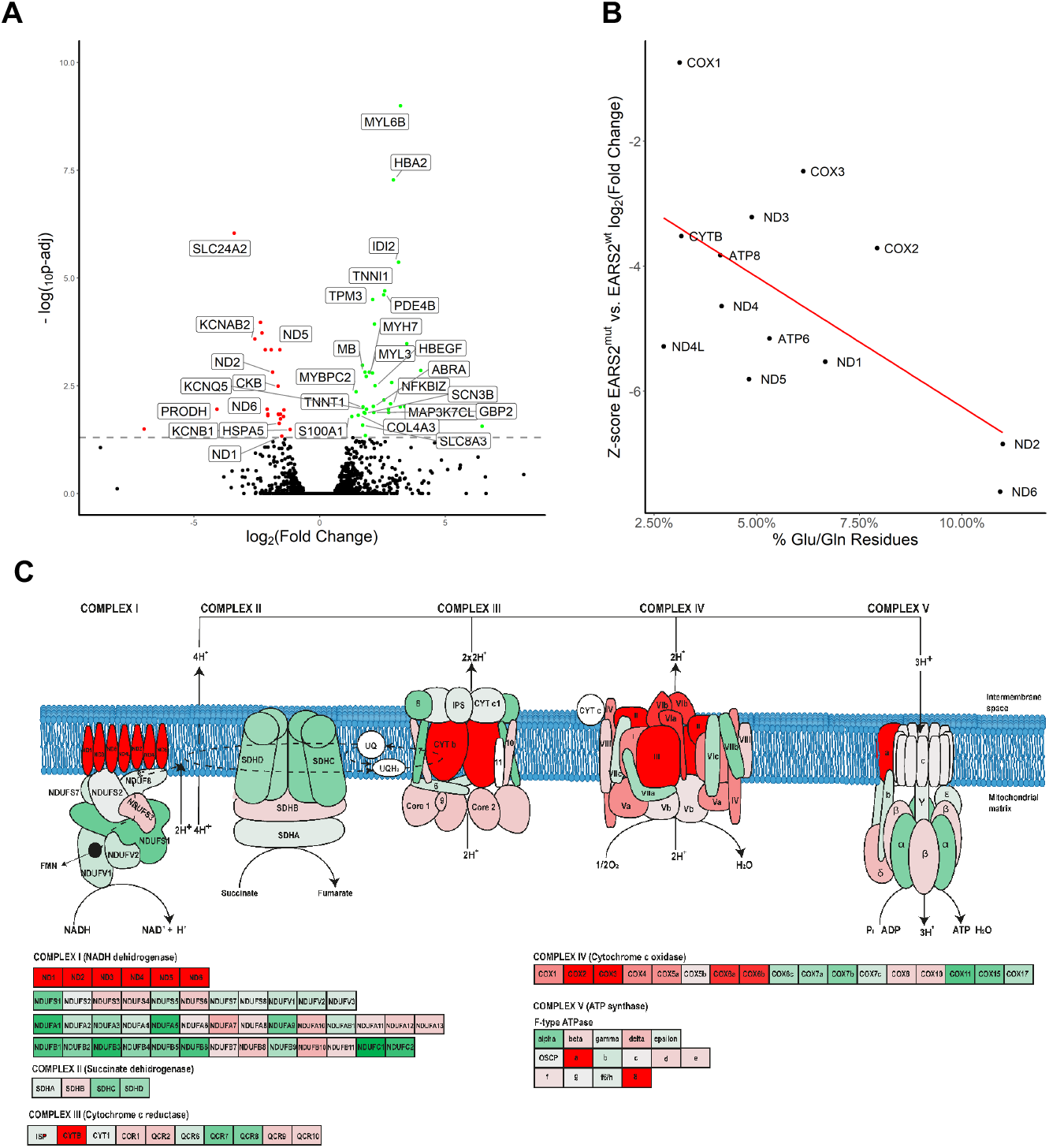
Differential gene expression of RIRCD muscle biopsies with digenic mutations (m.14674T>C plus heterozygous *EARS2)* compared to RIRCD muscle (m.14674T>C), without *EARS2* mutations. (A) Volcano plot analysis of transcriptomic changes where decreased genes are represented in red while the up-regulated ones are in green while genes that were not statistically significant are represented in black; (B) Analysis of altered mitochondrial gene expression resulted from our RNAseq showing that affected genes and their coding proteins carry a high numbers of glutamic acid and glutamine residues, in accordance to the mitochondrial translational defect; (C) Schematic representation of the mitochondrial respiratory chain enzymes resulted from the differential gene expression where red colour means significantly downregulated, green means significantly upregulated genes.

### Transcriptomic and proteomic changes in skeletal muscle biopsies identify a metabolic shift and increased mitochondrial biogenesis

We studied gene expression in the skeletal muscle of 6 RIRCD patients biopsied during the symptomatic phase and 6 healthy age-matched controls. Two of the RIRCD patients carried digenic mutations in *EARS2* (**Table 1**). Differential expression analysis in RIRCD biopsies identified 1398 RNAs with significant (p-adjusted ≤ 0.05) overexpression, and 1905 RNAs with significant under-expression compared to control biopsies (**Figure 1A-C**). We also analysed consecutive biopsies from 1 patient during the affected phase and after recovery, indicating metabolic alterations in RIRCD, which reversed in parallel with the clinical recovery **(Figure 1D)**. Comparative proteomic analysis of muscle protein extracts derived from 3 RIRCD patients **(Table 1)** versus 3 controls (3 months, 18 years, 19 years) led to the identification of 1600 proteins from which 141 were statistically significantly up-regulated and 134 down-regulated (p-Anova ≤ 0.05) in RIRCD muscle (**Figure 1E-F**). Changes in mtDNA encoded subunits and key proteins identified in our analysis were confirmed by immunoblotting (**Figure 2A-F)**. Integration of transcriptomic (*genes - italic*) and proteomic (proteins - normal) data was performed to highlight metabolic pathways, involved in the molecular mechanism of RIRCD.

Fibroblast Growth Factor 21 (*FGF21*), a hormone with crucial role in regulating lipid and glucose metabolism (Forsstrom, Jackson et al., 2019) and *GDF15*, both biomarkers of mitochondrial translation deficiencies were profoundly increased in RIRCD muscle and normalized after recovery (**Figure 1D**). Whether *FGF21* and *GDF15* are drivers of the observed metabolic shift in RIRCD patients, or just by-standing biomarkers is currently unclear. An increase in pyruvate dehydrogenase kinase isozyme 2 (PDK2), SUCLG2 and *ALDH6A* suggests a redirection of the metabolic flux from the respiratory chain to TCA, suggesting an increase in this metabolic pathway (Majer, Popov et al., 1998) and further supported by limited conversion of TCA components into glucose, illustrated by decreased PCK2 (Park, Jeon et al., 2018). Enzymes involved in the oxidation of fatty acids were up-regulated in RIRCD muscle (AC ADM, ACADS, ACAA2, ACAAT9), including enzymes that use short (ACSS1), medium and long chain fatty acids (ACOT9) in order to contribute to the acetyl-CoA pool, which is further channelled into the TCA (Forster & Staib, 1992, Kiema, Harijan et al., 2014, Tillander, Arvidsson Nordstrom et al., 2014). In addition, ACAT1 was decreased in patient muscle, slowing the conversion of acetyl-CoA into aceto-acetyl CoA and CoA, thus maintaining the cellular pool of acetyl-CoA needed for the TCA (Haapalainen, Merilainen et al., 2007). We also detected an increase in genes and proteins associated within serine and one carbon metabolism (*PHGDH, SHMT1*, SHMT2) **(Supplementary data)**, similar to changes reported in other mitochondrial myopathies (Forsstrom et al., 2019). All of this changes show that we have a strong metabolic shift from OXPHOS towards fatty acid oxidation and TCA cycle. We hypothesise that this metabolic remodelling acts as a compensatory mechanism in order to overcome the energetic deficit caused by blocked oxidative phosphorylation.

In addition, in our omics analysis we detected changes consistent with an activation of mTOR (mammalian target of rapamycin) and PI3K/AKT (phosphatidylinositol 3-kinase) in patient muscle (**Figure 1C**). To confirm the changes highlighted by altered expression of genes and proteins, we performed targeted immunoblotting for key players of these pathways. We detected an increase in S^2448^mTOR in affected and recovered muscle **(Figure 2C-D)**, while mTOR was increased just in the affected muscle, a decrease in *DEPTOR*, a known natural inhibitor of mTOR (Peterson, Laplante et al., 2009) and enhanced expression of *PIK3R3, PIK3CD* and *PIK3CG* in both affected and recovered muscle. Moreover, we saw an increase in PGC1α **(Figure 2C-D)**, a common target of mTOR which controls mitochondrial biogenesis (Cunningham, Rodgers et al., 2007), illustrated here, by a significant increase of mtDNA copy numbers at recovery **(Figure 2G)**.

### Amino acid sensing regulation of the mitochondrial respiratory chain may provide a muscle-specific rescue mechanism in RIRCD

Our data revealed that several pathways involved in the catabolism of amino-acids are increased in patient muscle (**Figure 1**). We observed a significant increase in GCN2, a serine/threonine kinase activated by amino-acid deprivation and S^51^eIF2α, a downstream target of GCN2 **(Figure 2E-F)**, in affected muscle. When phosphorylated, eIF2α stalls cap-dependent mRNA translation and initiates transcriptional stress response via ATF4, which was also increased in RIRCD muscle **(Figure 2E-F)** concomitant with stimulating the expression of amino acid biosynthetic and transporter genes (Masson, 2019).

An increase in proteins that act as transporters of different amino-acids (SLC38A1/*SLC38A1*, SLC1A7, SLC7A11/*SLC7A11*, SLC7A1/*SLC7A1*, TIM13, TIM50, TIM44, GRPE) (Kandasamy, Gyimesi et al., 2018) was observed in affected muscle. These proteins act as sensors for amino-acid status and are regulated in response to cellular needs. The increased need for amino acids in RIRCD muscle is further supported by notable increase in genes involved in the metabolism of alanine, aspartate and glutamate (*BCAT1, BACT2, FOLH1, ASPA, CAD, ASNS, DDO, ASS1, ASL*). The metabolism of branched chain amino-acids such as valine, isoleucine, lysine (DBT, GCDH, DGLUCY, ADHFE1) (Zhang, Zeng et al., 2017) and the degradation of lysine (GCDH) (Sauer, 2007) and tryptophan (AFMID/*AFMID*, KYNU/*KYNU*) (Pabarcus & Casida, 2002) were also significantly increased in RIRCD (**Supplementary data)**.

### Cysteine and glutamic acid depletion triggered combined respiratory chain deficiency in fibroblasts with m.14674T>C

Cysteine has been suggested to be an essential amino acid in the first months of life and its limited availability in infants may explain the reversible liver presentation of *TRMU* mutations (Zeharia et al., 2009) and potentially further compromises mt-tRNA^Glu^ steady state in RIRCD (Boczonadi, Smith et al., 2013). Glutamic acid is the cognate amino acid added by EARS2 to mt-tRNA^Glu^. Therefore, to understand if cysteine or glutamic acid availability influences mitochondrial translation in RIRCD we used primary fibroblasts of affected and unaffected mutation carriers of m.14674T>C and controls. These fibroblasts did not show respiratory chain deficiency in regular medium (Boczonadi, Smith et al., 2013). Lower cysteine levels in the cell culture medium (0.02 mM cysteine) for 12 days had a deleterious effect on complex I and complex IV in all fibroblasts, with an enhanced effect in cells from patients carrying the m.14674T>C mutation (**Figure 3A-B**). In contrast glutamine and glutamic acid depletion for 12 days triggered a mild, but significant decrease in MTCO1 and MTCO2 (Complex IV) **(Figure 3C-D)** in fibroblasts of patients with digenic heterozygous *EARS2* mutations. The observed changes were less severe than after cysteine depletion as glutamine and glutamic acid are considered to be non-essential and easily synthetized via the TCA (Liaw & Eisenberg, 1994).

Aminoacylation of mt-tRNA^Glu^ was not significantly altered in fibroblasts with digenic mutations compared to healthy homoplasmic carriers of m.14674T>C (**Supplementary Figure 5)**, while, studying aminoacylation in skeletal muscle was not possible due to the limited size of the biopsies.

**Figure 5.**
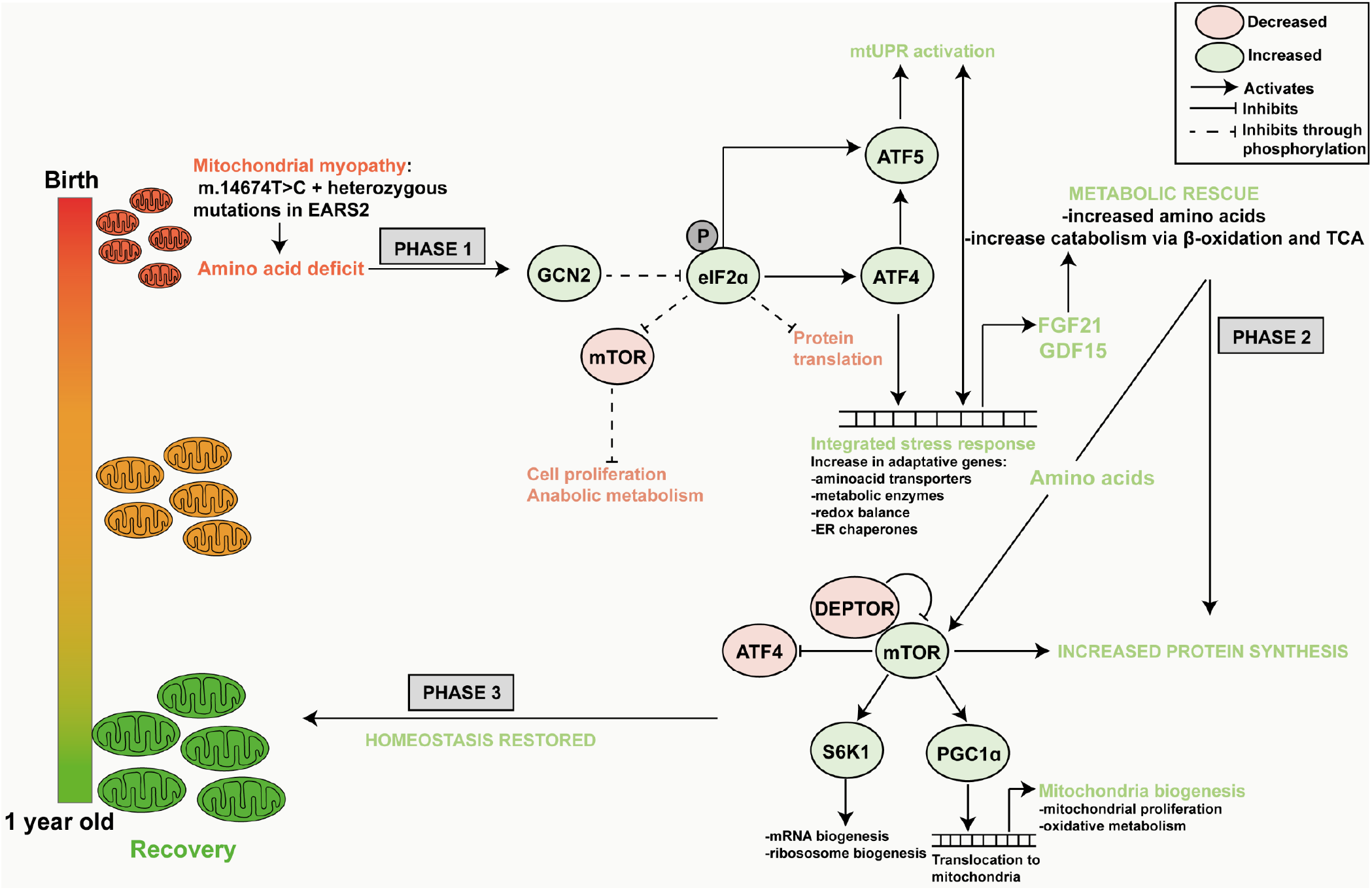
Graphical summary of changes identified in RIRCD muscle. Schematic representation of the metabolic changes in RIRCD. We propose a mechanism that takes place in three phases. Phase 1 consists in *FGF21* increase concomitant with the activation of the integrated stress via the GCN2-phospho eIF2A-ATF4 axis response which has a protective effect leading to increased expression of proteins involved in amino-acid transport and offers a metabolic rescue by increasing lipid oxidation and TCA. The second phase consists of mTOR activation which leads to increased mitochondrial biogenesis and protein synthesis and in phase 3 to recovery.

### Digenic mutations in m.14674T>C and *EARS2* result in a more severe defect of mtDNA-encoded gene expression in muscle

We compared differential expression of 2 RIRCD patients with digenic *EARS2* mutations (F2/1M, F5/1M) to 2 RIRCD patients without digenic *EARS2* mutations (F2/1M, F5/1M) and identified significantly altered (p-adjusted <0.05) gene expression of 59 mRNAs (36 increased, 23 decreased) including complex I proteins such as *ND1, ND2, ND4L, ND5, ND6* (**Figure 4A-C**). Skeletal muscle of RIRCD patients with digenic *EARS2* mutations showed lower expression of mtDNA-encoded genes. Furthermore, the degree of decrease in mRNA counts correlates directly with the proportion of glutamine and glutamic acid residues of the mtDNA-encoded proteins (r= 0.61677, p=0.024), with most severely affected subunit being NADH dehydrogenase, which contains the highest number of these amino acids (**Figure 4B**).

## Discussion

We studied 42 individuals from 19 families homoplasmic for m.14674T>C, and in 24 out of 27 affected individuals we identified additional heterozygous damaging variants in *EARS2* and in four other nuclear genes modifying or aminoacylating mt-tRNA^Glu^ (*TRMU, QRSL1, GOT2, GLS*). These variants were absent in healthy carriers of m.14674T>C. Three of the 9 *EARS2* variants detected in RIRCD patients were previously reported in patients with autosomal recessive leukoencephalopathy with thalamus and brainstem involvement and high lactate (LTBL) (Steenweg et al., 2012). While heterozygous pathogenic *EARS2* mutations do not lead to clinical symptoms, the co-occurrence of these variants with m.14674T>C in infants was associated with RIRCD (**Table 1**), and their disruptive effect on gene expression was demonstrated in skeletal muscle biopsies (**Figure 4**). The catalytic function of *EARS2* is to aminoacylate mt-tRNA^Glu^ at the 3’ end (m.14674T>C) (Steenweg et al., 2012), therefore the co-occurrence of these variants may further compromise the aminoacylation and the steady state of mt-tRNA^Glu^ leading to a digenic inheritance in RIRCD. However, fibroblasts carrying digenic mutations (m.14674T>C and *EARS2*) did not show significantly reduced aminoacylation of mt-tRNA^Glu^, compared to fibroblasts of healthy carriers of m.14674T>C (**Supplementary Figure 5**). It is possible, that aminoacylation of mt-tRNA^Glu^ would be more affected in skeletal muscle, however we could not study this in frozen biopsies. Additionally, EARS2 can have also non-canonical functions (e.g. amino acid sensing and regulation) thus the heterozygous mutations seen in RIRCD patients could alter these and worsen the effect of m.14674T>C. The other co-segregating nuclear variants may also affect aminoacylation of mt-tRNA^Glu^ and mt-tRNA^Gln^ (*QRSL1)* or glutamic acid and glutamine levels (*GOT2, GLS)* in mitochondria (**Supplementary Table 3)**. Nine RIRCD patients carried a common *TRMU* variant (p.Ala10Ser), previously shown to interact with m.1555A>G to cause deafness (Meng et al., 2017). *TRMU* downregulation in cells together with m.14674T>C alters mt-tRNA^Glu^ steady state (Boczonadi et al., 2013), suggesting again a digenic mechanism if these variants co-occur. In one family the index patient carried a variant in *MSS51* (Bareth, Nikolov et al., 2016, Garcia-Villegas et al., 2017), a protein that regulates translation and assembly of cytochrome *c* oxidase (**Table 1**). The prevalence of the co-segregating nuclear mutations may explain the model of only 1 out of ∼100 m.14674T>C mutation carriers expressing symptoms (**Supplementary Table 3**). True digenic inheritance has been reported in some other genetic conditions (facio-scapulo-humeral muscular dystrophy - FSHD, kidney diseases, deafness, etc.) (Nagara, Papagregoriou et al., 2018), however RIRCD is the first example of a digenic mtDNA-nuclear DNA interaction.

We previously reported that >30% of normal mt-tRNA^Glu^ steady state in healthy carriers of homoplasmic m.14674T>C is sufficient to maintain mitochondrial protein synthesis in skeletal muscle, while individuals <15-20% mt-tRNA^Glu^ were affected (Horvath et al., 2009). Here we show that the co-occurrence of heterozygous *EARS2* (or other nuclear variants with similar function) further compromise the fragile compensatory rescue mechanisms in infants and act as a second hit, explaining the clinical manifestation of RIRCD. Transcriptomic and proteomic signature of RIRCD skeletal muscle biopsies detected profoundly increased *FGF21* and *GDF15* and significant changes in several components signalling a metabolic shift from OXPHOS towards lipid metabolism and the TCA cycle, while glycolysis was reduced (**Figure 1)**. In support of our findings, increased CK, low carnitine, slightly increased acyl-carnitines, increased TCA cycle metabolites and accumulation of lipids, glycogen and abnormal mitochondria have been reported previously in RIRCD patients (Boczonadi et al., 2015).

*FGF21* has been recently described to drive the integrated mitochondrial stress response and activate a cascade of events in mitochondrial myopathy patients and mice, thus leading to a distinct metabolic remodelling (AlJohani, Khan et al., 2019, Forsstrom et al., 2019, Lehtonen, Forsstrom et al., 2016, von Holstein-Rathlou, BonDurant et al., 2016). Here we show, that *FGF21* is associated with the activation of an integrated stress response (increased GCN2, phospho EIF2α, ATF4 and ATF5) (Pakos-Zebrucka, Koryga et al., 2016) in RIRCD, concomitant with an increase in fatty acid oxidation and TCA cycle associated enzymes, thus, leading to an alternative energy source (bypassing complex IV and V) which, most likely to compensate for the lack of ATP. In contrast to progressive mitochondrial myopathies that have a long term ISR, RIRCD patients present with a milder short term ISR (significant decrease of GCN2, ATF4 and eIF2A in recovered muscle (**Figure 2E-F)** which induces a beneficial hormetic response by activating protective cellular mechanisms that protect the cells against amino-acid deprivation, metabolic insults and oxidative stress(Pakos-Zebrucka et al., 2016).

Our data suggest that the events in RIRCD muscle take place in three distinct phases **(Figure 5)**. In phase 1 the limitation of amino acids results in activation of the mitochondrial integrated stress response (ISR), with the activation of GCN2, which phosphorylates Eif2α at Ser^51^ that, inhibits general cytosolic protein synthesis and increases the translation of specific proteins such as ATF4 and ATF5 **(Figure 5)**, which control the transcription of numerous adaptive genes that are involved in maintaining the cellular homeostasis (amino-acid transporters, metabolic enzymes, ER chaperones) (Melber & Haynes, 2018, Xia, Lei et al., 2018). This in turn helps to conserve nutrients thus providing a temporary relief from metabolic stress (Ron, 2002). In phase 2, after the metabolic crisis is surpassed, we have an activation of mTOR in RIRCD muscle supported by decreased levels of DEPTOR and increased levels of S^2448^mTOR, S6K1 kinase and PGC1α. This leads to increased mitochondrial biogenesis and elevation of mt-tRNA^Glu^ steady state, resolving the block in mitochondrial translation in phase 3 (Cunningham et al., 2007).

The up-regulation of TCA in recovering infants further contributes to the metabolic salvage and facilitates the synthesis of amino acids. In support of our findings, a recent study demonstrated that in a mouse model of abnormal mitochondrial translation (*Mrps12*^*ep/ep*^) there is a similar mitochondrial stress response that orchestrates the metabolic rescue by increasing TCA together with an increase in cell proliferation and mitochondrial biogenesis(Ferreira, Perks et al., 2019).

The skeletal muscle is the largest reservoir of amino acids, particularly glutamate and cysteine which are structural components of skeletal muscle, explaining the muscle specific presentation of RIRCD (Owen, Kalhan et al., 2002). Glutamate is also the most abundant amino acid in breast milk and its concentration increases in the first 4 months of breastfeeding, suggesting the imminent need for this amino acid in infants (Koletzko, 2018). Although we do not have metabolomic data regarding the levels of amino acids in our patients, the significant changes in genes and proteins involved in amino acid metabolism highlight their importance in RIRCD **(Figure 5)** (Forsstrom et al., 2019, Owen et al., 2002). We investigated the effect of amino acid deprivation (cysteine, glutamic acid, and glutamine) in RIRCD fibroblasts and detected respiratory chain deficiency **(Figure 3)**. We showed previously that the respiratory chain defect in RIRCD myoblasts can be rescued by cysteine supplementation, by improving cysteine dependent thiourydilation, and detected reduced level of thiourydilation in skeletal muscle, highlighting that amino acids may regulate oxidative phosphorylation (Boczonadi et al., 2013). Furthermore, supplementation with cysteine rescued the defect of mitochondrial protein synthesis in fibroblasts carrying mutations in *TRMU, MTO1* or mt-tRNAs (m.3243A>G, m.8344A>G) (Bartsakoulia, Mueller et al., 2016). This might suggest that the cysteine-dependent thiouridylation of the mt-tRNA^Glu^ catalyzed by *TRMU* is impaired, supported by the detection of *TRMU* variants in 9 of the 27 in RIRCD patients.

The clinical recovery is characteristic for RIRCD, however the importance of sufficient supply of amino acids is likely to be relevant in other mitochondrial myopathies such as m.3243A>G or mtDNA deletions. Further studies are needed to explore the role of potentially targetable amino acid sensing pathways and their impact in mitochondrial diseases.

In summary, we show that RIRCD is a skeletal muscle specific disease, where digenic mutations in mitochondrial and nuclear DNA interact with reduced nutritional intake of amino acids and activate a cascade of metabolic events including integrated stress response activation, FGF21 signalling, mTORC1 activation, metabolic shift to TCA and fatty acid oxidation and enhanced mitochondrial biogenesis. This stress response in turn improves the availability of amino acids (glutamate, cysteine) which facilitates the metabolic shift, which in turn improves mt-tRNA^Glu^ steady state, and leads to recovery. Similar digenic nuclear-mitochondrial interactions may be relevant in other human mtDNA related diseases.

## Materials and methods

### Experimental models and ethics approval Ethical approval

The human fibroblasts were obtained from the Biobank of the MRC Neuromuscular Translational Research Centre (London-Cambridge-Newcastle). All the necessary ethical approvals are available in these facilities (REC reference 08/H0906/28+5). Human skeletal muscle was obtained by biopsy. Patients and their family members have given consent to the study “Genotype and phenotype in inherited neurological diseases” study (REC: 13/YH/0310, end date 30/09/2023, IRAS ID: 2042290) which includes all relevant permission for this work. The consent form contains also the consent for any omics analysis.

### Human skeletal muscle

*Vastus lateralis* muscle needed for this study was collected from patients via needle biopsy. Collected samples were frozen in liquid nitrogen and stored at -80°C. Sample codes used for the RNA sequencing are as follows: F2/1 M, F2/2 F, F3/1 M, F3/2 M, F4/1 F, F5/1M, F6/1 M. Samples used for proteomics and western blotting are as follows: F2/1 M, F2/2 F, F3/1 M, F3/2 M. All controls were sex/ age matched. All details related gender, age and other additional information can be found in Table 1.

### Cell lines

The human primary fibroblasts were obtained from the Newcastle Biobank from donors via skin biopsy utilizing routine protocols. The samples were obtained from the following patients: Patient (F7/7F-healthy carrier, F7/2M-RIRCD, F7/5F-healthy carrier, F7/3F-RIRCD without a clear second mutation. For more details relating the patients see Table 1. HEK293T cells (+/+) and *PDE12* knock-out cells (-/-) and -/- cells expressing WT *PDE12*, p.Glu351Ala (catalytic mutant), Δ16 (coding for *PDE12* lacking 16 first aa), Δ23 mutants (coding for *PDE12* lacking 23 first aa), and p.Arg23Trp *PDE12* Cdna and Human 143B osteosarcoma (HOS) were obtained from Michal Minczuk.

### Whole exome sequencing

We performed whole exome sequencing in 34 individuals, homoplasmic for m.14674T>C and bioinformatics analysis, as described previously(Griffin et al., 2014). Variants that were annotated as splice-site, frameshift or stop-loss/gain and also non-synonymous variants that were predicted to be damaging, show evolutionary conservation and were present in a known InterPro domain were considered potentially pathogenic and were interrogated for their predicted in silico deleteriousness and previous known association with human disease as described (Richards, Aziz et al., 2015, Taylor, Pyle et al., 2014). Variants were filtered for those that were exclusively present in either affected or unaffected individuals. Expected population minor allele frequencies were obtained from the Human Mitochondrial DataBase (http://www.hmtdb.uniba.it/hmdb/) and the Exome Aggregation Consortium (http://exac.broadinstitute.org/). The NIHR BioResource, UK BioBank and 1000 Genomes datasets were queried for m.14674T>C/G mutation carriers.

### Cell culture

Primary human fibroblasts from controls and patients were grown in high glucose Dulbecco’s modified Eagle’s medium (DMEM, Thermo Fisher Scientific 11965084) supplemented with 2 mM L-glutamine (Thermo Fisher Scientific 35050061) and 10% foetal bovine serum (Thermo Fisher Scientific 26140) at 37°C, in a humidified 5% CO_2_ atmosphere. Primary fibroblasts were left to grow in normal conditions and then split between normal and cysteine and glutamine depleted medium.

For the cysteine depletion experiment cells were incubated in cysteine free medium which was purchased from Thermo Fisher Scientific (21013 DMEM, high glucose, no glutamine, no methionine, no cysteine). Cell culture medium was supplemented with Glutamine (Thermo Fisher Scientific 35050061, 2 mM), Sodium Pyruvate (Thermo Fisher Scientific 11360070, 0.11 mg/ml), 5% dialyzed FBS (Thermo Fisher Scientific, A3382001) and 0.02 mM of L-cysteine (Sigma Aldrich C7352) which represents 10% of the standard DMEM concentration, which was enough to support cell growth. Cell were incubated for 12 days in the cysteine depleted media at 37°C, in a humidified 5% CO_2_ atmosphere.

For the Glutamine depletion experiment cell were grown in standard high glucose DMEM with no glutamine added (Thermo Fisher Scientific 11965084) supplemented with 5% dialyzed FBS (Thermo Fisher Scientific, A3382001) for 12 days at 37°C, in a humidified 5% CO_2_ atmosphere.

Cell lines to study PDE12 were as cultured as described in Pearce et al. 2017(Pearce et al., 2017). HEK293 knock-out cell line expressing the cDNA coding for p.Arg23Trp was constructed also as described previously by Pearce et al. 2017. Briefly, HEK293 Flp-In T-Rex was purchased from Invitrogen-R78007 which allows for the generation of stable, doxycycline-inducible expression of transgenes by FLP recombinase mediated integration. This system was used to generate the PDE12-/-cell lines which inducible expressed PDE12.Strep2.Flag (PDE12.FST2) and the mutants p.Glu351Ala, Δ16, Δ23 mutants and p.Arg23Trp *PDE12* cDNA. To generate the PDE12 -/- and PDE12+/-cell lines, HEK293 were transiently transfected to express a pair of CompoZr ZNFs (Sigma Aldrich), which targeted the *PDE12* gene locus at exon 1. Cells were then electroporated with pZFN1 and pZFN2 using Cell Line Nucleofector (Lonza) and buffer kit V (Lonza VVCA-1003) applying program A-023. 72 hours post transfection single-cells were sequenced by Sanger sequencing to identify clones harbouring indels in *PDE12*.

PDE12 cells and Human 143B osteosarcoma (HOS) cell lines were cultured in DMEM (Thermo Fisher Scientific 11965084) containing 2 mM Glutamax (Thermo Fisher Scientific 35050061), 10% FBS (Thermo Fisher Scientific 26140), Penicilin/Streptomycin (Gibco 15140122) at 37°C, in a humidified 5% CO_2_ atmosphere.

### RNA sequencing

RNA was extracted with the mirVana miRNA isolation kit according to the manufactures protocol (Thermo Fisher Scientific AM1560). RNAseq libraries were prepared with Illumina TruSeq Stranded (Illumina MRZH11124) polyA enriched RNA with Ribo-Zero Human (Illumina 20020594) kit and were sequenced on Ilumina HiSeq 2500 Platform according to the paired-end protocol, as previously described(Burns, Donkervoort et al., 2018). The quality of sequencing reads was checked with FastQC. Raw sequence reads were trimmed to 50bp with FastX-toolkit v.0.0.14(http://hannonlab.cshl.edu/fastx_toolkit/index.html) and aligned to complete Human (hg38) reference genomes, using the STAR aligner v.2.5.3a two-pass protocol that is outlined in the GATK documentation(Dobin, Davis et al., 2013). Number of reads mapped to Ensmbl genes was counted with HTSeq v.0.9.1(Anders, Pyl et al., 2015) and the differentially expressed genes were identified using the DESeq2 v.1.12.4 package(Love, Huber et al., 2014).

### Quantification of mitochondrial DNA copy number

The relative mtDNA copy number per cell was quantified by a multiplex Taqman (Bio-rad 4369510) qPCR assay by amplifying *MT-ND1* (mitochondrial encoded gene) and *B2M* (nuclear encoded gene) with a CFX96™ Real-Time PCR Detection System (Bio-Rad) following the protocol described previously(Bartsakoulia et al., 2016). The primers used for template generation of standard curves and the qPCR reaction are as follows B2M: Fw-CACTGAAAAAGATGAGTATGCC, Rv-AACATTCCCTGACAATCCC; MTND1: Fw-AACATTCCCTGACAATCCC, Rv-AACATTCCCTGACAATCCC. The copies per µL of each template were standardized to 1 * 10^10^ and a serial dilution in 1Log_10_ dilution steps, was amplified along with the DNA negative control on each qPCR plate. This was performed in 20 µL reactions in a 96 well-plate (Bio-Rad 5496), sealed using microplate ‘B’ plate sealers (VWR 391-1293). The reaction mixture was composed of: 5 µL × 5 × Taqman (Bio-rad 4369510), 0.4 µM of reverse and forward primers, 25-50 ng of DNA template, 0.2 µL MyTaq HS DNA polymerase (Bioline BIO-21112) and PCR-grade autoclaved sterile deionised water (to make up 20 µL reaction mixture). The cycling conditions were as follows: 1) initial denaturation at 95°C for 3 minutes, 2) 40 cycles of denaturation at 95°C for 10 seconds, 3) annealing and extension at 62.5°C for 1 minute. The relative mtDNA copy number was calculated using the ΔCt data following the eqation: CopyNumber = 2 (2^-ΔCt^) where Delta Ct (ΔCt) equals the sample Ct of the mitochondrial gene (MTND1) subtracted from the sample Ct of the nuclear reference gene (B2M).

### Aminoacylation of mt-tRNA^Glu^ and mt-tRNA^Gln^

The impact of identified variants on the aminoacylation of mt-tRNA^Glu^ was analyzed using RNA isolated from fibroblasts grown in standard DMEM, and in fibroblasts grown in MEM with 1% the amino acid concentration present standard DMEM for 48 hours. RNA was extracted from sub-confluent fibroblasts using Trizol (Thermo Fisher Scientific 15596026) according to the manufacture’s protocol, and the final RNA pellet was dissolved in 10◻mM sodium acetate (Sigma Aldrich S2889-250G), pH 5.0 at 4◻°C. For the deacylated (dAc) control, the pellet was resuspended in 200◻mM Tris-HCl (Applichem Biochemica A3452) at pH 9.5 and incubated at 75◻°C for 5◻min, followed by RNA precipitation and resuspension in 10◻mM sodium acetate (Sigma-Aldrich S2889-250G) buffer pH 5. Next, 5◻µg of RNA was separated on a 6.5% polyacrylamide gel (Thermo Fisher Scientific NP0321BOX) (19:1 acrylamide:bisacrylamide) containing 8◻M urea in 0.1◻M sodium acetate pH 5.0 at 4◻°C and blotted to Hybond N+ membranes (Amersham RPN303B). Following UV-crosslinking, the blots were washed in hybridization buffer of 7% SDS (Carl Roth CN30.1), 0.25 M sodium phosphate (Sigma-Aldrich 342483-500G) pH 7.6 for 1◻hour at 65◻°C. The membrane was subsequently incubated overnight at 65◻°C in hybridization solution with ^32^P-labeled antisense RNA probes (Hartman Analytic), generated by in vitro transcription by T7 RNA polymerase in the presence of ^32^P-labeled alpha-UTP (Hartman Analytic), using linearized templates. After hybridization, the blots were washed six times with 1x SSC (Sigma-Aldrich S6639) for 15◻min at 65◻°C. Bound probes were detected by phosphor imaging on an Amersham Typhoon Scanner.

### Protein lysate preparation and immunoblotting analysis

Patient (F2/1M, F3/1M, F3/2M, F5/1M) and control derived muscle(1 month, 17 years old female and 17 years old male) were lysed in 100 µL of lysis buffer (50 mM Tris-HCl-Applichem Biochemica A3452 (pH 7.8) 150 mM NaCl (Merck 1064041000), 1 % SDS (Carl Roth CN30.1), and Complete Mini-Roche 11873580001) using a manual glass grinder. Control and patient fibroblasts were lysed in RIPA buffer (Sigma-Aldrich R0278) with Complete Mini by pipetting. Then samples were centrifuged for 5 min at 4°C and 5000 *g*. Protein concentration of the supernatant was determined by BCA assay (ThermoFisher 23225) (according to the manufacturer’s protocol).

For immunoblot studies, 10 μg protein was used in each case, loaded on a gradient polyacrylamide gel (NuPage 4-12% Bis-Tris Protein gels ThermoFisher WG1402BOX, NP0321BOX) and separated for 120 min at 120 V. Following the separation, proteins were transferred to PVDF membrane (Iblot2 Transfer stacks IB23001) using the iBlot2 system (ThermoFisher IB21001) according to the manufactures protocol. Membranes were blocked with 5% Milk prepared in PBS-T for 2 h followed by four washing steps using PBS (Gibco 18912014) with 0.1% Tween20 (Sigma-Aldrich P7949-100ML) (PBS-T). Membranes were incubated with several primary antibodies (Table 1) at 4 °C (overnight) and then washed in PBS-T thrice. Horseradish peroxidase conjugated secondary goat anti-rabbit antibody (ThermoFisher Scientific 31460) or goat anti-mouse antibody (ThermoFisher Scientific 31430) was diluted at 1:25,000 and added to membranes for 1 h. Next, membranes were washed three times in PBS-T for 10 min. By using the enhanced chemiluminiscence, horseradish peroxidase substrate (Super-Signal West Pico 34577 and Super-Signal West Femto 34094; Pierce) signals were detected using a UVItech machine.

### Comparative proteomic analysis

#### Cell lysis, sample preparation and trypsin digestion

In total seven muscle samples (*vastus lateralis)* derived from three healthy controls and four patients were processed independently. Approximately 10 slices of 10µm of muscle were lysed in 50 µL of lysis buffer (50 mM Tris-HCl (Applichem Biochemica A3452) (pH 7.8) 150 mM NaCl, 1 % SDS (Carl Roth CN30.1), and Complete Mini Roche 11873580001) using a manual glass grinder. Then samples were centrifuged for 5 min at 4°C and 5000 *g*. Protein concentration of the supernatant was determined by BCA assay (ThermoFisher 23225) (according to the manufacturer’s protocol) and cysteines were reduced with 10 mM of DTT (Roche 10708984001) by incubation at 56°C for 30 min. Next, the free thiol groups were alkylated with 30 mM IAA (Sigma-Aldrich I1149-25G) at room temperature (RT) in the dark for 30. Sample digestion and cleanup were performed using filter-aided sample preparation (FASP) as described previously(Roos, Preusse et al., 2019) with some minor changes: 100 µg of protein lysate was diluted 10-fold with freshly prepared 8 M urea/100 mM Tris-HCl (Applichem Biochemica A3452) (pH 8.5) buffer and placed on PALL microsep centrifugal device(Merck Z648051) (30 KDa cutoff) and centrifuged at 13,500 *g* at RT for 20 min (all the following centrifugation steps were performed under the same conditions). Three washing steps were carried out with 100 µL of 8 M urea (Sigma Aldrich U1250)/100 mM Tris-HCl (Applichem Biochemica A3452) (pH 8.5). For buffer exchange, the device was washed thrice with 100 µL of 50 mM NH_4_HCO_3_ (pH 7.8) (Sigma-Aldrich S2889-250G). The digestion buffer contains as follows (final volume of 100 µL): trypsin (Promega V5117) (1:25 w/w, protease to substrate), 0.2 M GuHCl (Sigma-Aldrich G3272-500G) and 2 mM CaCl_2_ (Sigma-Aldrich C3306) in 50 mM NH_4_HCO_3_ (pH 7.8) (Sigma-Aldrich S2889-250G), which was added to the concentrated proteins and the samples were incubated at 37°C for 14 h. Resulting tryptic peptides were recovered by centrifugation with 50 µL of 50 mM NH_4_HCO_3_ (Sigma-Aldrich S2889-250G) followed by 50 µL of ultra-pure water. Afterwards, the resulting peptides were acidified (pH<3 by addition of 10 % TFA (v/v) (Biosolve 213141). All digests were quality controlled as described previously.

#### LC-MS/MS analysis

Samples were measured using an Ultimate 3000 nano RSLC system coupled to an Orbitrap Fusion Lumos mass spectrometer (both Thermo Scientific). Peptides were preconcentrated on a 100 µm × 2 cm C18 trapping column for 10 min using 0.1 % TFA (v/v) at a flow rate of 20 µL/min. Next the separation of the peptides was performed on a 75 µm × 50 cm C18 main column (both Pepmap, Thermo Scientific 164567) with a 120 min LC gradient ranging from 3-35 % of 84 % ACN (Biosolve 12041), 0.1 % FA (v/v) (Biosolve 69141) at a flow rate of 230 nL/min. MS^1^ spectra was acquired in the Orbitrap from 300 to 1500 m/z at a resolution of 120000 using the polysiloxane ion at *m/z* 445.12003 as lock mass(Olsen, de Godoy et al., 2005),with maximum injection times of 50 ms and ACG target was set at 2.0×10^5^ ions. Top fifteen most intense signals were selected for fragmentation by HCD with a collision energy of 30 %. MS^2^ spectra were acquired in the ion trap at a resolution of 120,000, with maximum injection times of 300 ms, a dynamic exclusion of 15 s. The ACG target was set at 2.0×10^3^ for MS^2^.

#### Label free data analysis

Data analysis of the acquired label free MS data was performed using the Progenesis LC-MS software from Nonlinear Dynamics (Newcastle upon Tyne, U.K.). Raw MS data was aligned by Progenesis which automatically selected one of the LC-MS files as reference. After automatic peak picking, only features within retention time and *m/z* windows from 0-120 min and 300-1500 m/z, with charge states +2, +3, and +4 were considered for peptide statistics and analysis of variance (ANOVA) and MS/MS spectra were exported as peak lists. Peak lists were searched against a concatenated target/decoy version of the human Uniprot database (downloaded on 22.07.2015 containing 20273 target sequences) using Mascot 2.4 (Matrix Science, Boston, MA, USA), MS-GF+, X!Tandem and MyriMatch with the help of searchGUI 3.2.5(Vaudel, Barsnes et al., 2011). Trypsin was selected as enzyme with a maximum of two missed cleavages, carbamidomethylation of Cys was set as fixed and oxidation of Met was selected as variable modification. MS and MS/MS tolerances were set to 10 p.p.m and 0.5 Da, respectively.

To obtain peptide-spectrum match and to maximize the number of identified peptides and proteins at a given quality we used PeptideShaker software 1.4.0 (http://code.google.com/p/peptide-shaker/). Combined search results were filtered at a false discovery rate (FDR) of 1 % on the peptide and protein level and exported using the PeptideShaker features that allow direct re-import of the quality-controlled data into Progenesis. Peptide sequences containing oxidized Met were excluded from further analysis. Only proteins that were quantified with unique peptides were exported. For each protein, average of the normalized abundances (obtained from Progenesis) from the analyses was calculated in order to determine the ratios between the patient muscle and control.

Only proteins which were (*i*) commonly quantified in all the replicates with (*ii*) unique peptides, (*iii*) an ANOVA p-value of ≤0.05 (Progenesis) and (*iv*) an average ratio ≤ log_2_ -2.2 or ≥ log_2_ 0.98 were considered as up respectively down regulated. AThe Proteomap was generated using the online available tool (https://www.proteomaps.net/). The annotation of these proteomaps is based on the KEGG database platform, each protein is shown by a polygon, and functionally relevant proteins are arranged as neighbors. Additionally, polygon areas represent protein abundances weighted by protein size.

#### Quantification and statistical analysis

Data was plotted using GraphPad Prism v.7.0 software (GraphPad Software, USA) or Origin 6.0 (Origin Lab) and Adobe Illustrator Artwork 23.0 (Adobe Systems). The statistical test and method is indicated in the legend of the figures. P-values of less than 0.05 were considered statistical significant for all experiments.

## Data Availability

Genomic data is available via RD-CONNECT while proteomics and transcriptomic data submission to Gene Expression Omnibus (Geo)I and ProteomeExchange is still in progress

## Data and code availability

All the data generated or analysed during this study is included in this article or in the supplemental methods and are available from the corresponding author upon request. The exome sequencing data is made available also via RD Connect while the RNA-seq and Proteomics data is deposited on Gene Expression Omnibus data Repository or ProteomeExchange respectively. All the code used for the exome sequencing and RNA-seq is available via Bioconductor or GitHub.

## Acknowledgments

We thank the NIHR BioResource volunteers for their participation, and gratefully acknowledge NIHR BioResource centres, NHS Trusts and staff for their contribution. We thank the National Institute for Health Research and NHS Blood and Transplant. The views expressed are those of the author(s) and not necessarily those of the NHS, the NIHR or the Department of Health. The analysis of allele frequencies in the UK Biobank Resource were conducted under application number 18794. We thank Reinaldo Issado Takata and Alessandra de la Rocque Ferreira for molecular studies and Dr. Luisa Iommarini (University of Bologna) for the fruitful discussion about the cysteine depletion experiment.

## Author contributions

HG, MJJ, WW, CC and VM performed bioinformatics analysis. HG, MJJ, MG and EPH were involved in RNA sequencing. DH and AR completed the proteomics studies. DH, MG, CP, SFP and VB performed cell culture experiments. BM studied mtDNA copy numbers, AP, JD, and BM performed Sanger sequencing. JP, US, PDSP, MH, KJ, AC, JFP, MMN, AM, KC, SS, JU, MH, MT, SDM and RH contributed patients to the study. AGD, VM, HL, MM and PFC contributed constructive comments on bioinformatic analysis and interpretation of data. HG, DH, JSM and RH were involved in the design of the experiments, data analysis and writing of the manuscript

